# Efficient and Practical Framework for Bias Estimation in Spectral CT

**DOI:** 10.64898/2026.03.11.26346993

**Authors:** Olivia F. Sandvold, Roland Proksa, Amy E. Perkins, Peter B. Noël

## Abstract

**Background:** Spectral computed tomography (CT) is increasingly used for quantitative imaging, yet accurate prediction of spectral quantitative bias remains challenging and computationally expensive with conventional approaches. Bias manifests as systematic deviations in reconstructed quantities (e.g., Hounsfield units, iodine density) from their true physical values. It arises from a combination of model mismatch, hardware/processing imperfections, exam-dependent factors, and noise-induced effects amplified by nonlinear operations such as the logarithmic transformation and material decomposition.

**Purpose:** We present a practical, projection-based statistical framework to estimate noise-induced spectral bias efficiently, without the runtime burden of Monte Carlo (MC) simulation.

**Methods:** To demonstrate the bias estimator, we modeled the central-ray of a clinical X-ray tube attenuating through a 300 mm patient-equivalent path with a 10 mm insert containing 10 mg/mL iodine. A 120 kVp tube voltage and tube currents from 100-350 mA were used. Ideal and realistic photon-counting detector responses were simulated across 50 bin threshold settings. Quantum Poisson noise was modeled, and Bayesian probabilities of material decomposition outputs centered on ground truth iodine and water bases were computed. Expected material decomposition outputs 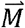 were derived from a 2D probability map, and bias was measured. A simple Python Monte Carlo (MC) simulation served as a reference.

**Results:** The proposed bias estimator closely matched MC-derived bias, with an average relative iodine bias percent difference between the estimators of 0.44% across all tube currents and bin thresholds. Average runtime of the bias estimator was only 0.5% of the MC simulation. Optimal thresholds for minimizing iodine noise (via the Cramér-Rao lower bound) differed from those minimizing iodine bias, highlighting key noise-bias tradeoffs.

**Conclusion:** Efficient spectral bias and noise estimation are essential for quantitative CT system design. This modular framework enables rapid, bias-aware optimization of spectral acquisition parameters and is adaptable to alternative spectral CT technologies beyond photon counting.

**Novelty and Significance of Study:** *Please briefly (150 words or less) describe the novelty and/or significance of your study*.

Bias estimation is paramount for designing accurate spectral CT systems that deliver improved diagnostic performance. Traditional approaches rely on computationally intensive Monte Carlo simulations. We propose an efficient and practical bias estimator that uses Bayesian statistics and expected material decomposition values derived from a flexible, modular CT forward model. Unlike conventional Monte Carlo approaches, this framework enables rapid exploration of spectral design tradeoffs between bias and noise. We demonstrate both the accuracy and speed of this bias estimator relative to Monte Carlo approaches.

## 1. Introduction

In computed tomography (CT), accurate modeling of both qualitative image appearance and quantitative performance is essential to maximize diagnostic benefit for patients. Manufacturers use these metrics to optimize system design parameters, set scan protocols, and predict improvements relative to previous technologies. With nearly half of CT scans in the US acquired with iodinated contrast agents^1^ and evidence showing spectral CT imaging increases differentiation efficacy^2,3^ and reduces radiation dose^4,5^, precise estimation of spectral quantitative bias has become a critical priority. This bias manifests as systematic errors in reconstructed quantities, such as Hounsfield units or iodine density, causing them to deviate from their true physical values. Quantitative bias can be caused by systematic errors, including physical imperfections (e.g., hardware inconsistencies, scatter) and software/processing errors (e.g., spectral model mismatch). Additionally, bias can originate from examination-related sources (e.g., patient motion, mispositioning). Bias is also induced by stochastic X-ray quantum processes, resulting in noise-induced bias^6^. Signal noise is transformed non-linearly due to the application of the logarithm function–a necessary operation performed on energy-integrated intensities or discrete photon counts to translate X-ray information into projection line integrals. In the presence of X-ray Poisson or electronic noise, the mean output may be skewed from the expected output of the log transform. This bias amplification process becomes especially significant in the clinical setting of low-dose acquisitions. We propose a practical statistical analysis framework designed to estimate the extent bias in spectral domains.

Because bias correction methods can be difficult to implement robustly in routine clinical workflows, often requiring stable calibration^7,8^, accurate system modeling^9^, and high computational burden, bias reduction through optimized acquisition and processing offers a practical alternative. By selecting system settings and processing strategies that intrinsically limit bias (e.g., appropriate thresholds, exposure levels, etc.), one can achieve meaningful improvements in quantitative accuracy without adding substantial complexity or computational overhead. This is accomplished by modeling X-ray attenuation and computing reconstruction effects in output projections or images. Approaches to modeling bias include Monte Carlo (MC) simulations^10,11^ and convex optimization methods based on Taylor series expansions^11,12^. These bias estimators are often computationally expensive, which limits rapid prototyping/system optimization. Additionally, most approaches to bias modeling have focused solely on conventional attenuation results, neglecting spectral imaging and subsequent bias magnification in spectrally derived images (e.g., iodine density maps, virtual non-contrast images). This technological gap also hinders the task-specific design and optimization of spectral CT systems for clinical applications.

The Cramér-Rao lower bound (CRLB) is a common and popular method to estimate the minimum theoretical noise within a CT system and is easily extendable into spectral domains^13,14^. It is essential for CT optimization but cannot accurately describe noise-induced bias due to the assumption of an unbiased estimator. Thus, we introduce a projection-based, bias-aware model that is flexible and readily adaptable to a wide range of spectral CT acquisition instrumentations (e.g., source or detector-based spectral imaging).

## 2. Methods

This Methods section outlines the framework used to evaluate spectral CT imaging performance. We begin by outlining the X-ray emission spectrum and the detector characteristics relevant to this study, followed by a phantom model designed to emulate a realistic clinical imaging scenario. Next, we present our novel statistical estimator for bias. Finally, we evaluate its performance relative to a Monte Carlo and CRLB simulation.

### 2.1 X-ray emission spectrum

The incident X-ray emission spectrum was modeled from a tungsten anode source (vMRC800, Philips Healthcare) in units of number of photons per mAs per keV per steradian using SpekCalc software^15–17^. Tube housing attenuation and aluminum filtration were applied. The X-ray tube spectrum was denoted as *S*(*E*). The tube voltage was fixed at 120 kVp. The tube current was initialized at 100 mA and then varied from 100 to 350 mA in 50 mA increments to study the impact of dose on bias estimator performance. In this paper, *E* ranged from 10 to 120 keV in 1 keV increments. These tube settings replicated clinical protocols and ensured sufficient photon counts (i.e., no starvation events) for the tested patient size (Section 2.3). The central ray of the X-ray beam was modeled as a one-dimensional projection-based simulation to demonstrate the bias estimator. However, our method can be readily applied to the full cone beam geometry. This pencil-beam simplification isolates noise-induced spectral bias independent of geometric reconstruction effects.

### 2.2 Detector characteristics

The spectral sensitivity of a photon-counting detector (PCD) was modeled using pulse-height spectrum *H*(*P,E*)describing the probability that an incoming photon with energy *E* generates an electronic pulse with amplitude *P*. For an ideal PCD, the detector response is perfectly energy resolving, 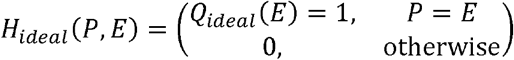 The limited stopping power of a practical direct-conversion layer can be incorporated if the thickness *d*_DC_and attenuation μ _*DC*_ (*E*)are known, 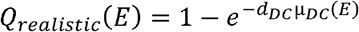. Real PCDs deviate from the ideal response due to effects including charge sharing, electronic noise, incomplete charge collection, and pulse pileup. These mechanisms broaden the pulse-height spectra and reduce energy resolution. The spectral sensitivity of a bin spanning thresholds (*T* _*low*_, *T*_*high*_) is given by 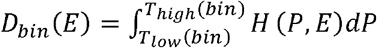 and represents the integration of all pulse heights within the bin threshold range.

A prototype cadmium zinc telluride (CZT) PCD with two bins was modeled using experimentally measured pulse-height spectra that include detector imperfections. In addition, the spectral stopping power of the direct-conversion CZT sensor material was incorporated. These measurements were performed at the European Synchrotron Radiation Facility (ESRF) in 2014. They are used as representative realistic detectors rather than implementing a specific commercial system. For all simulations, we used an electronic noise-excluding bin threshold level at 20 keV. **Figure 1A-C** demonstrates realistic spectral bin overlap contained in the modeled system spectral responses,, with varying bin threshold values of 45, 70, and 90 keV compared to an ideal detector response with a bin threshold at 70 keV (**Fig. 1D**).

**Figure 1:**
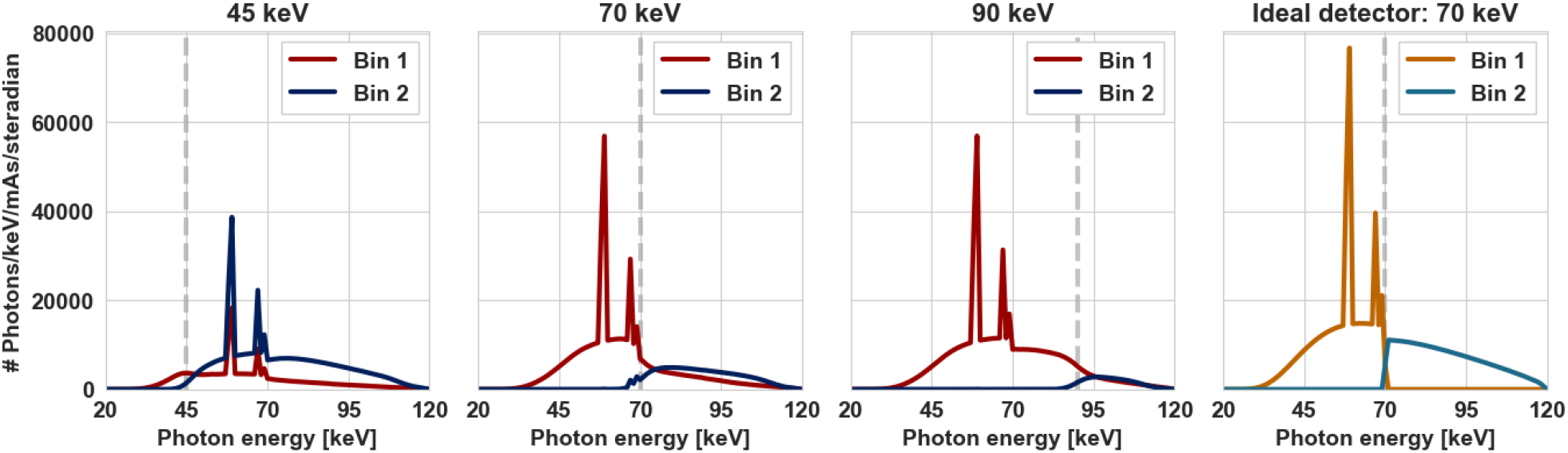
Simulated PCD system responses using the incident 120 kVp, 100 mA tube spectrum. The grey dashed vertical line represents the defined threshold between bins for each configuration. **A-C** demonstrate realistic PCD bin overlap, while **D** depicts an ideal bin spectral separation at 70 keV.

Changing the bin threshold not only affects the system spectral separation but also influences the balance of the number of photons contained across the bins. To investigate these effects on spectral bias, we varied the bin threshold level from 50 to 110 keV in 50 equidistant steps. While varying the bin threshold level, we held the tube current constant at 100 mA.

### 2.3 Phantom model

To mimic a clinically relevant scenario, we modeled a 300 mm diameter water [1 g/mL] cylinder. We inserted an additional 10 mm diameter cylinder containing 10 mg/mL iodine to recapitulate a medium-sized vessel carrying iodinated contrast. Energy-dependent attenuation characteristics were sourced from NIST for pure iodine and water^18^. A single pencil-beam X-ray projection through the phantom center was simulated for the study.

### 2.4 Practical spectral decomposition bias estimator

To estimate spectral material bias, we first define a two-dimensional material parameter space

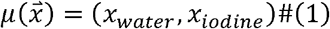

corresponding to water and iodine basis materials respectively. All material parameters are expressed in g/mm^2^. For consistency, all variables with an arrow represent a 2D vector. The measurement space is a two-dimensional photon-count space

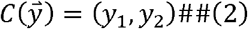

representing the counts recorded in the two energy bins described by:

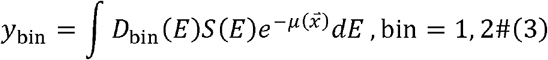

A generally nonlinear forward model

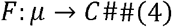

maps material parameters to 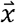 expected photon counts 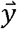. For a given ground-truth vector 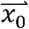, the corresponding expected counts are

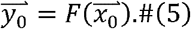

A bounded region Ω ⊂ μ is defined around 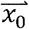 such that it contains most material estimates expected from noisy measurement decomposition. The limits of Ω are determined by first computing the basis material-specific CRLB variances 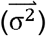 as described by Roessl and Hermann^14^. For iodine and water individually, the bounds of Ω are set as 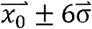 to capture > 99.9% of possible obtainable material path lengths. The region Ω is discretized into equidistant number (*n*) of material samples. For each pair of samples, 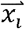, belonging in Ω ranges, the forward model is evaluated to obtain detector counts

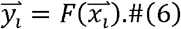

Assuming a known independent Poisson noise model for the photon counts, the detection probability is computed as

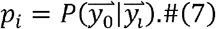

Since the probabilities, *p*_*i*_, are defined in count space, they must be transformed back into material space. This is achieved by using a Jacobian normalization of the forward model. Let

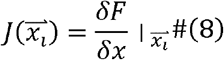

denote the Jacobian matrix evaluated at 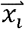

The non-normalized material-space probability weights are given by:

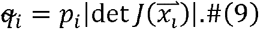

After normalization, the discrete material-space probability distribution becomes

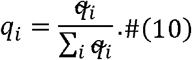

This *q*_*i*_, represents the 2D probability mass function over the range of material path lengths (Ω, *n* x *n*)The expected material estimate is computed as the first moment of this distribution:

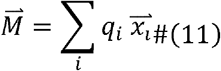

where 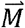. represents the expected material decomposition values (*M_water_, M_iodine_*) of the posterior distribution given the probability space *q*_*i*_ generated from the set of samples 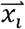 The bias in the material space is then defined as

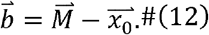

The Jacobian normalization ensures conversion of probability mass under the nonlinear transformation between material space and count space and is essential for obtaining an unbiased estimate in the presence of model nonlinearity. **Equation 13** expresses the relative material decomposition bias % error for iodine and water. This metric is used to compare varying system configurations and to estimate the lowest theoretical relative bias in each material domain after decomposition.

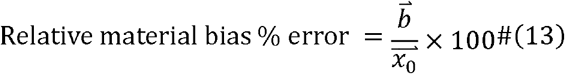

For each system configuration and simulation, the total runtime in seconds was computed from steps following **Equation 5** to **Equation 12** using the Python timeit library.

### 2.5 Monte Carlo simulation validation

We designed a basic Python Monte Carlo (MC) simulation to validate the bias estimator. Expected bin counts and variance were computed as defined in **Equation 3**. We tested six tube currents (100, 150, 200, 250, 300, 350 mA) and two bin threshold levels (45, 70 keV). A curve-fitting maximum likelihood solver was implemented for solving the two basis function outputs (in water and iodine) for each bin-count pair. The Python SciPy library minimize function was used with the Broyden–Fletcher–Goldfarb–Shanno algorithm to minimize the negative log likelihood between randomized counts and detector responses.

To ensure sufficient MC sample size, *N*, for each system configuration at each threshold and dose level, we ran pilot MC simulations using *N*_*p*_ independent Numpy random Poisson values for each bin. *N*_*p*_ varied from 5,000 to 40,000 in steps of 1000. **Equation 14** describes the process to select *N*: either when the MC signal-to-noise ratio (SNR) measured in the iodine channel is greater than or equal to 10 or maximally to 40,000 if the SNR condition is never met. This upper limit was set to avoid excessive computational time at the cost of the presence of greater MC variance in addition to intrinsic material decomposition variance in the system. We predict that low bias acquisitions will require larger *N* compared to cases with greater inherent spectral bias.

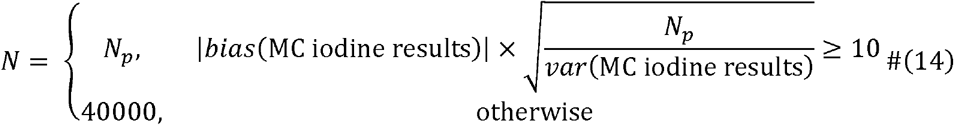

To account for variability produced in each MC simulation, ten repeats of the MC estimator were used for each system configuration with *N* determined sample points. The total MC estimator runtime, which included noise generation, material decomposition, and bias estimation, was measured in seconds using the Python timeit library. The mean bias and the standard deviation of bias estimates across the ten repetitions were computed.

## 3. Results

### 3.1 Bias estimator using bin threshold=45 keV

We illustrate the results of the bias estimator using a 120 kVp, 100 mA acquisition with a with bin threshold set at 45 keV. **Figure 2A** shows the 2D normalized *q*_*i*_ probability map generated the bounded region Ω for water and iodine plotted on the x- and y-axes, respectively. Dashed yellow lines mark 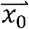. The magnitude of *q*_*i*_ is shown with the color gradient expressed in the color bar. **Figure 2B** contains *q*_*i*_ values ≥ 10^−5^ to better visualize the shape of the estimator. **Figs. 2C** and **2D** show the marginal probability of estimates for each basis material. The material estimate *M*. was located at (299.02, 0.16) [g/mm^2^] water/iodine in contrast to 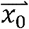 located at (300,0.1) [g/mm^2^]. This point, marked by green stars, represents the weighted probability center of mass, or the expected material outcome for the sampled values. Finally, **Fig. 2E-F** highlights the estimated water and iodine bias, 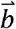 or the difference between 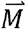 and 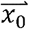. The relative bias error for 10 mm of 10 mg/mL iodine and 300 mm of 1 g/mL water was 56.9% for iodine and −0.3% for water, given the defined system settings.

**Figure 2.**
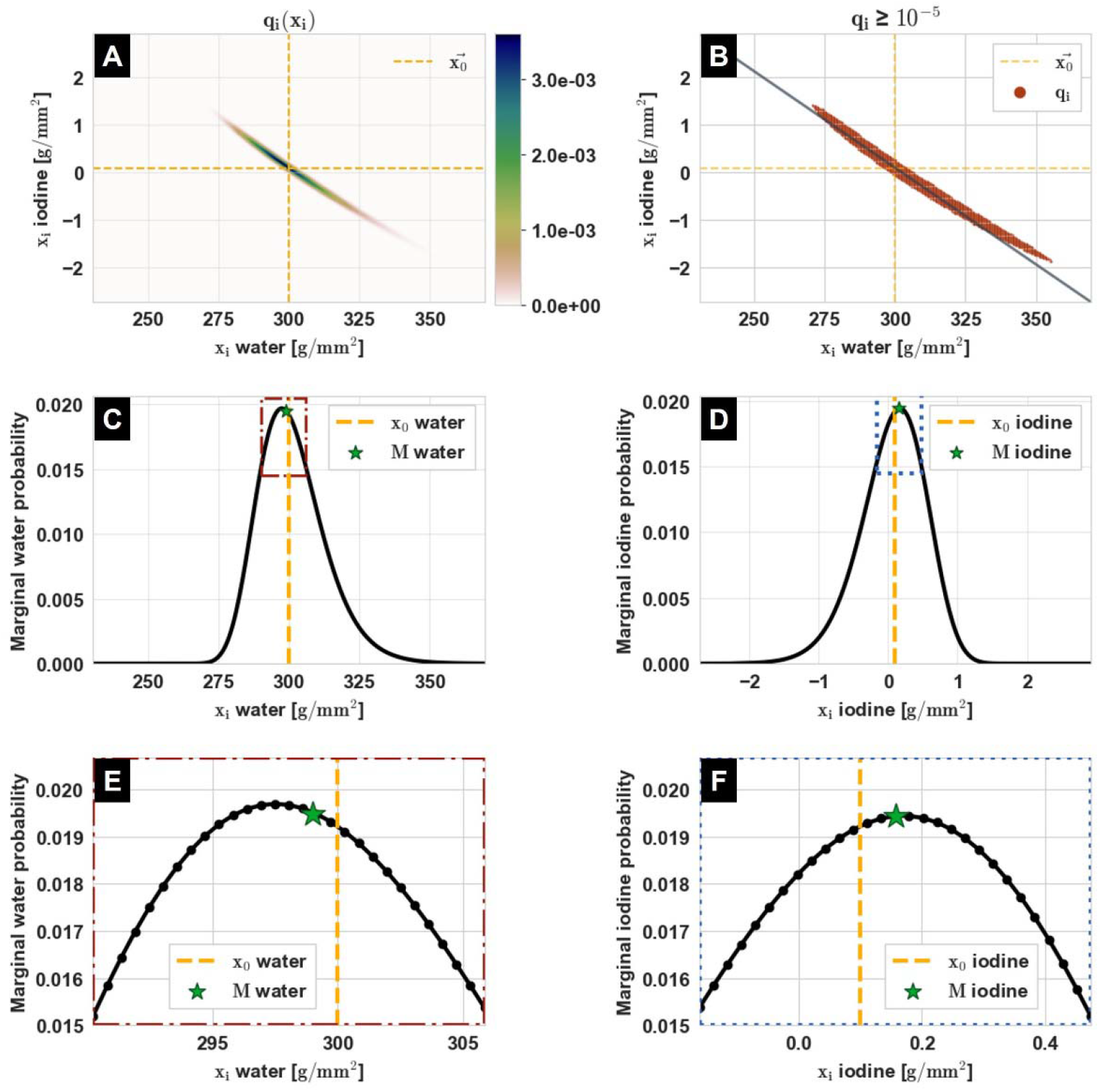
(A) Normalized 2D Bayesian probability map over Ω-bounded 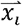 values. The yellow dashed lines denote 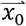 (B) 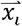 points with normalized probability q_i_ ≥ 10^−5^, with the solid gray line indicating the negative linear iodine-water decomposition trend through 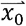. (C–D) Marginal probability distributions for water and iodine decomposition estimates, illustrating their non-normal, skewed shapes. **M** for each material is marked with the green star. (E–F) Marginal probability density for water and iodine path length estimates for probabilities ≥ 0.015.

The 2D probability map (**Fig. 2A-B**) featured a strong anticorrelated, non-linear relationship between the basis function outputs. For water underestimates, iodine overestimates were more probable. In the individual marginal material estimation probability plots (**Fig. 2C-D**), a skewed, non-normal distribution was given. This matched with expectations given multiple nonlinear effects: applying the logarithm to noisy data and the nonlinear mapping from photon counts to the iodine–water material decomposition basis further amplifying this shift.

### 3.2 Bias estimation performance at varying bin threshold levels

The 2D probability maps were generated at varying bin thresholds and for both realistic and ideal PCDs, and the bias values were estimated (**Fig. 3, Table I**). For iodine estimation, the bias % error was smallest using the ideal, perfect spectral separation case with an estimated iodine bias error of 0.9%. In the realistic 45 keV bin threshold system design, the estimated iodine bias was 56.9%, the poorest outcome of all tested bin threshold values. At this bin threshold, the estimated water % error was −0.3%, which was also the largest water error over the tested bin thresholds. All tested bin thresholds, regardless of detector response setting, had positive bias error estimates for iodine, with iodine error % at least forty times greater than the water error %.

**Figure 3.**
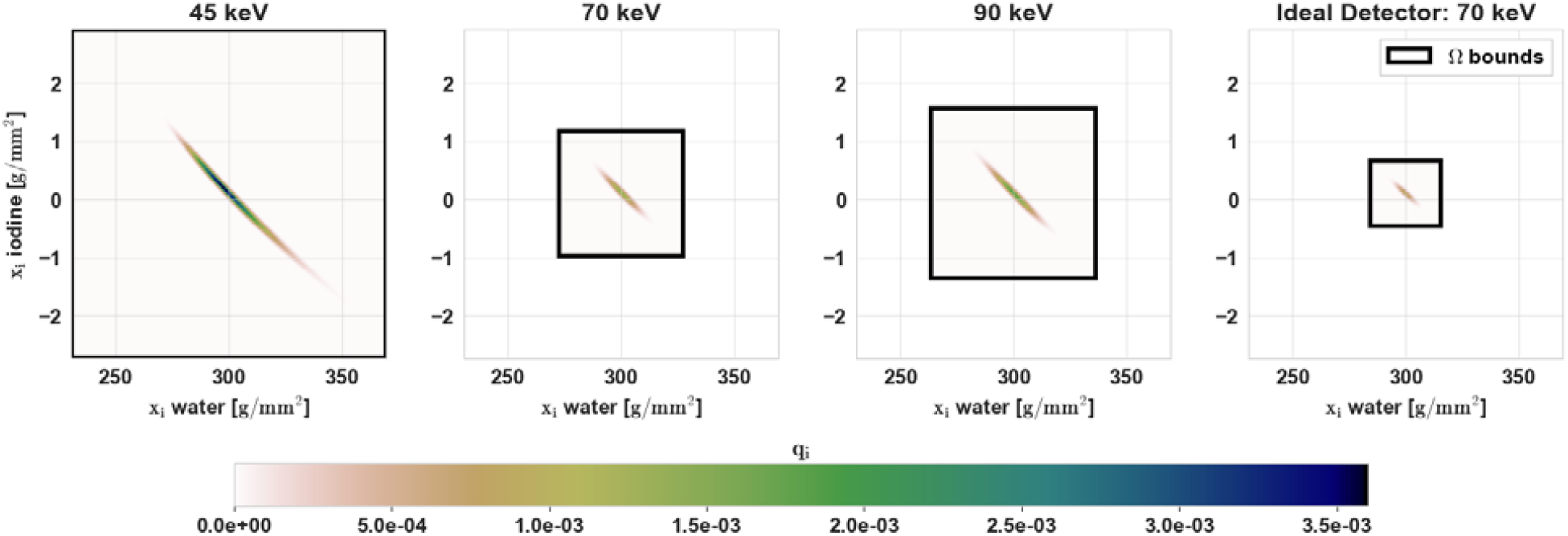
2D probability maps for three realistic PCDs using bin thresholds of 45, 70, and 90 keV (left) vs ideal PCD with bin threshold at 70 keV (right). The black box outlines each estimator’s range.

**Table I.** Bias comparison of realistic and ideal PCD responses at varying bin thresholds.

|  | Rel. bias error % |  | range [g/mm<sup>2</sup>] |  |
| --- | --- | --- | --- | --- |
|  | Water | Iodine | Water | Iodine |
| Realistic response<br>Bin threshold |  |  |  |  |
| **45 keV** | -0.327 | 56.9 | [230.6 – 369.4] | [-2.7 – 2.9] |
| **70 keV** | -0.001 | 2.9 | [272.4 – 327.6] | [-1.0 – 1.2] |
| **90 keV** | 0.028 | 1.2 | [263.5 – 336.5] | [-1.4 – 1.6] |
| **Ideal response**<br>**Bin threshold** |  |  |  |  |
| **70 keV** | 0.002 | 0.9 | [284.5 – 315.5] | [-0.5 – 0.7] |

One primary characteristic of the bias estimator is the use of the CRLB to condition the search area of Ω bound 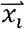 values. The 45 keV bin threshold with realistic PCD had the widest Ω range, followed by the 90 keV and 70 keV realistic configurations, and finally the 70 keV ideal PCD, in order of decreasing range.

### 3.3 Bias estimator compared to CRLB estimator

Both estimated biases and CRLB variances in the iodine and water domains were simultaneously produced. **Table II** contains comparisons of material-specific noise measured in *q*_*i*_ and the CRLB for the same system configurations. For all materials and thresholds tested, the bias estimator probability map noise (standard deviation, std) was greater than or equal to the CRLB noise. For the tested thresholds, the *q*_*i*_ water and iodine noise metrics on average were 0.8% and 1.5% greater than the respective CRLB estimates.

**Table II.** Noise estimated using bias estimator compared to CRLB noise.

| | Std of $q_i$ [g/mm<sup>2</sup>] | | CRLB noise [g/mm<sup>2</sup>] | |
| --- | --- | --- | --- | --- |
|  | Water | Iodine | Water | Iodine |
| **Realistic response**<br>**Bin threshold** |  |  |  |  |
| **45 keV** | 11.891 | 0.496 | 11.570 | 0.470 |
| **70 keV** | 4.610 | 0.180 | 4.603 | 0.179 |
| **90 keV** | 6.100 | 0.243 | 6.085 | 0.242 |
| **Ideal response**<br>**Bin threshold** |  |  |  |  |
| **70 keV** | 2.588 | 0.093 | 2.586 | 0.093 |

In **Figure 4**, the estimated bias error % was plotted over CRLB-estimated noise for water and iodine as the bin threshold levels varied.

**Figure 4.**
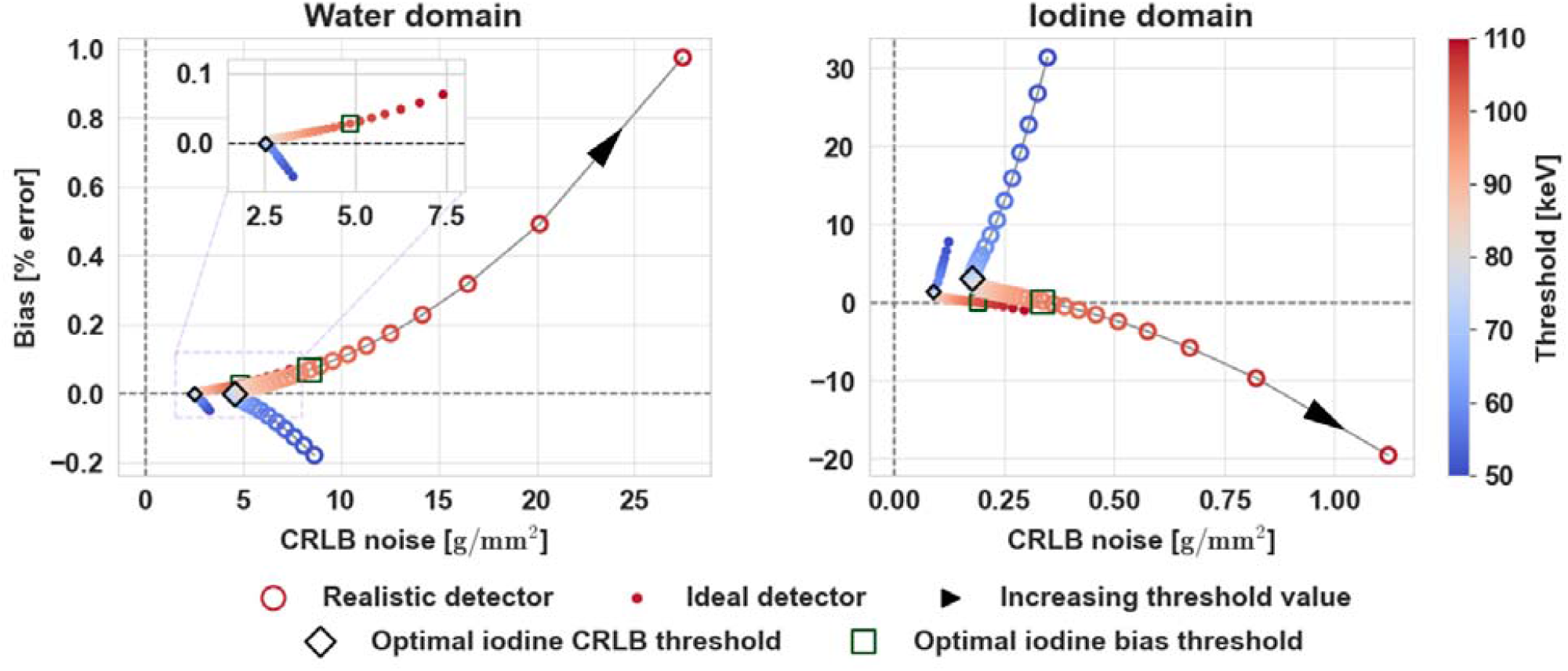
CRLB noise and bias % error for water (left) and iodine (right) material estimates as bin threshold values changed from 50 to 110 keV (dark blue to dark red). The results show performance using realistic PCD responses (open circles) compared to an ideal PCD (closed circles).

Increasing the bin threshold reduced the CRLB⍰estimated noise for both detector types and in both material channels, up to an inflection point—the minimum CRLB noise estimate threshold—corresponding to 64.7 keV for the ideal detector and 69.6 keV for the realistic detector. Material domain noise then rose with bin threshold following the optimal CRLB noise threshold. Water and iodine CRLB noise were minimized at the same threshold level.

In contrast, the optimal absolute iodine bias error percentages—0.11% for the realistic detector and −0.01% for the ideal detector—occurred at 99 keV and 102.7 keV, respectively. The iodine and water biases were not jointly optimized using the same threshold level. Water biases were minimized at bin thresholds of 70.8 keV using a realistic PCD and 67.1 keV for an ideal PCD. Across all thresholds, the ideal detector consistently produced lower CRLB noise and smaller absolute bias errors for both water and iodine compared with the realistic detector.

The threshold that minimized CRLB iodine noise was lower than the threshold that minimized iodine bias. With the realistic detector, using the threshold that minimized iodine bias resulted in a CRLB noise level 1.89x higher than the optimal (minimum⍰noise) CRLB value. The material decomposition bias followed an anticorrelated pattern: increasing negative water bias corresponded to increasing positive iodine bias, and vice versa. The relationship between noise and bias over the tested thresholds was nonlinear in both material channels.

### 3.4 Bias estimator and Monte Carlo comparison

Figure 5. shows the results of iodine SNR due to changing and the total runtime of the MC and bias estimator simulations for two different bin threshold levels: 45 keV (left) and 70 keV (right), over tube currents from 100 to 350 mA.

**Figure 5:**
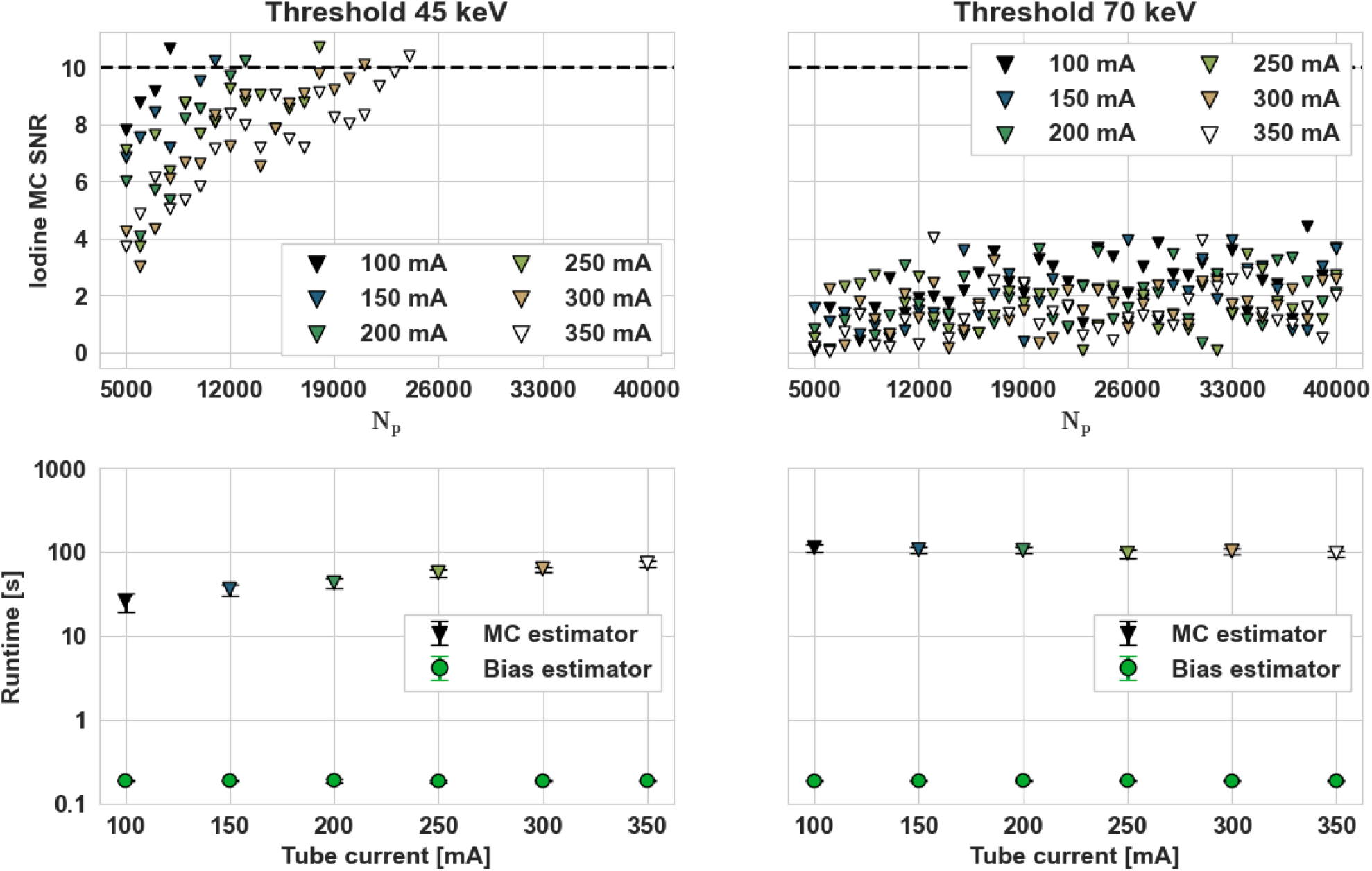
(Top row) MC SNR in iodine channel for each tube current as increased. (Bottom row) Computation runtime of the bias estimator compared to the mean Monte Carlo simulation runtime. Standard errors are plotted over ten repetitions of each estimator.

With the 45 keV bin, as the tube current increased, iodine bias decreased, necessitating larger *N* values to satisfy the SNR minimum requirement. In contrast, for the 70 keV bin threshold system, across all tested mA settings, the MC SNR in the iodine channel remained below 10. Therefore, *N* was set to 40,000 for all 70 keV MC simulations. Comparing runtime, the bias estimator consistently performed faster than the MC simulations. The bias estimator runtime was independent of the tube current. MC estimator runtime increased as tube current and *N* increased for the 45 keV threshold systems. Over all tested tube currents and defined bin thresholds, the bias estimator took an average of 0.5% or less of the total runtime of the MC simulation.

Figure 6. demonstrates that the bias estimator and mean MC simulation bias contained consistent iodine bias percent error agreement, with average absolute differences of 0.68% at the 45 keV threshold and 0.21% at the 70 keV threshold, across all tube currents. All bias estimator estimates were within one standard deviation of the Monte Carlo average output across the ten repetitions. Finally, both the bias estimator and Monte Carlo bias estimates followed a decreasing monotonic, inverse relationship with tube current. Fitting a minimized least-squares, inverse relationship function using both sets of bias estimates over all tested tube currents yielded R^2^ coefficients of 0.998 and 0.959 for the 45 and 70 keV bin threshold sets respectively.

**Figure 6.**
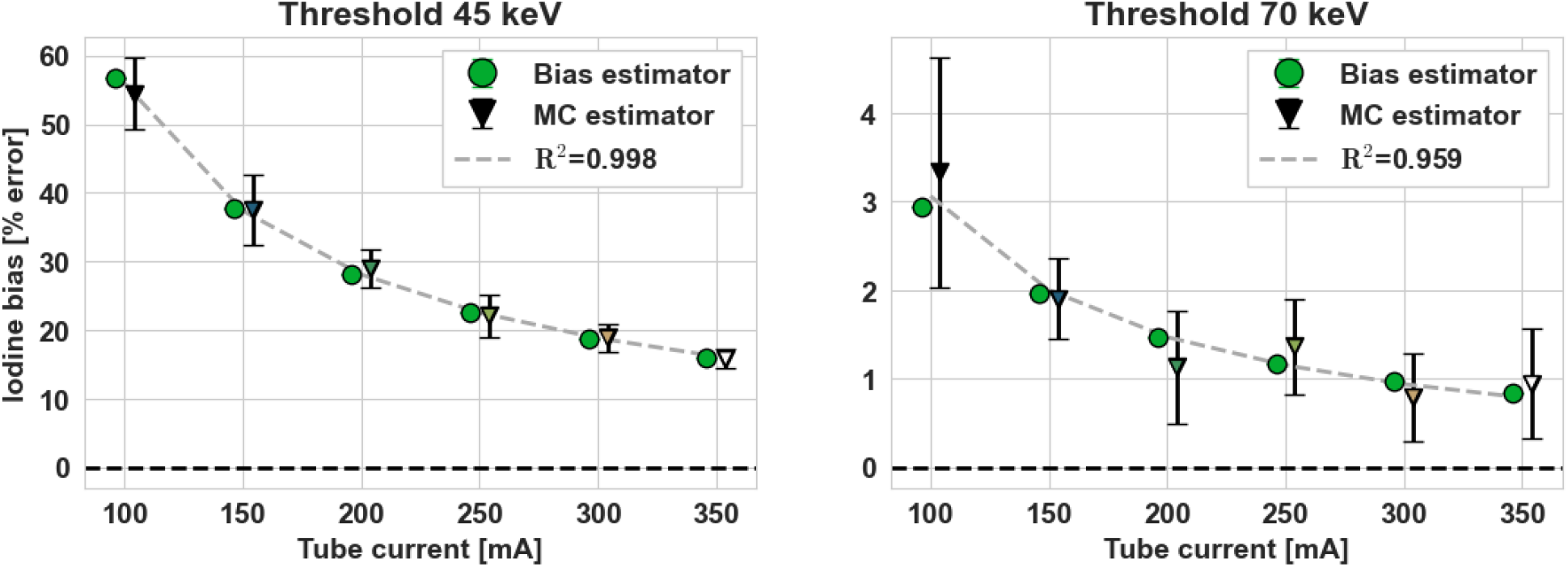
Estimated iodine bias % errors produced in the bias estimator compared to MC results. The mean MC bias over the ten runs is marked with a triangle, and the standard deviation of iodine bias over the runs is marked with error bars. For each threshold level, the least-squares, best fit inverse function using both estimators’ results is plotted with gray dashed line.

## 4. Discussion

The proposed bias estimator provides an efficient means of statistically characterizing spectral bias while avoiding the computational cost of Monte Carlo simulations. Because the framework is modular, system components can be exchanged to reflect a variety of realistic CT configurations, extending naturally beyond the photon-counting example shown here to rapid kVp-Switching, dual-layer detectors, and dual-source acquisitions by incorporating the related forward model and noise characteristics. The photoelectric effect and Compton scatter bases can be modeled to capture bias in downstream virtual non-contrast, virtual monoenergetic, or even physical density maps. Importantly, this framework enables bias-aware CT system and parameter optimization prior to hardware implementation. Our results demonstrate that selecting energy-bin thresholds to reduce estimator bias yields different bin positions than those obtained by optimizing solely for the minimum Cramér–Rao lower bound, highlighting that variance-optimal designs need not be bias-optimal in spectral CT. CRLB optimal settings prioritize balanced bin counts compared to low iodine bias acquisitions emphasizing high spectral separation. This highlights a key trade-off in system design and the value of having both an accurate bias model and a noise model. In many clinical settings, accurate quantification (e.g., vessel iodine concentration or attenuation) is more consequential than subtle differences in noise^19,20^. By providing direct estimates of systematic bias, our framework enables optimization decisions that explicitly prioritize quantitative fidelity, allowing acquisition and reconstruction parameters to be tuned toward minimizing clinically relevant bias rather than focusing exclusively on noise-related metrics.

Our projection-based analysis indicates that, for the material decomposition of the central ray alone, there exist energy-bin threshold settings in photon-counting CT (PCCT) at which the iodine decomposition bias can be driven to (approximately) zero. In a complete imaging chain, achieving truly zero bias is more challenging because additional systematic errors can arise from image reconstruction, regularization, calibration or model mismatch, and exam-specific factors (e.g., patient size and spectrum hardening). Notably, at the bin-threshold “zero-crossing” points, the corresponding CRLB noise for iodine is substantially higher—a 1.89x increase relative to the noise-optimal thresholds. We show that the noise of the statistical bias estimator is greater than or equal to the CRLB noise. However, for system settings that yield bias < 5%, the bias estimator noise remains within 1% of the CRLB-predicted material domain noise. Together, these findings highlight a fundamental bias-variance tradeoff in spectral CT design.

To provide a reference for 1D projection bias estimation, we implemented a baseline MC approach for material decomposition comparisons. The simulation models Poisson-distributed quantum noise and incorporates realistic detector effects, including charge sharing, pulse pileup, and finite stopping power. In the pilot study, we selected N such that the MC SNR for iodine was greater than or equal to 10 to ensure limited MC noise. However, this condition was not satisfied with any of the 70 keV threshold scenarios due to low iodine bias. Therefore, the MC variance has a larger impact on mean bias estimates which is evident when comparing R^2^ coefficients. These MC comparisons confirm that the proposed estimator provides reliable bias predictions across a range of conditions.

Numerous methods exist to model photon-counting detectors and quantify propagation of quantitative errors through the imaging chain. These include cascaded models^21^, use of pre-computed lookup tables^22^, statistical correlations of neighboring pixels^23^, and application of the maximum likelihood estimation with an expanded Taylor series^12^ to approximate the bias of a given CT system. While these approaches have been used to characterize attenuation-based data, direct estimation of bias in material decomposition outputs has not been widely explored. This work seeks to provide an accessible bias estimation tool for clinically relevant spectral imaging tasks. Across all CT modeling approaches, photon starvation remains a central challenge. Various approximations^24–28^ (e.g., metal artifact reduction, filtration, substitution) have been defined to address zero⍰count measurements; all methods affect downstream noise appearance, attenuation measurements, and image artifacts^29^. The proposed estimator would require specific knowledge of the implemented approach(es) in low photon conditions to predict changes in material decomposition bias. As clinical scenarios causing photon starvation (e.g., low dose acquisitions, large patient habitus, presence of metal implants) are unavoidable, the accurate modeling of system performance in these settings is crucial.

Recent CT development trends make bias-aware spectral modeling increasingly important. Clinical PCCT systems reduce electronic noise, enable tunable spectral separation, and support ultra⍰high⍰resolution imaging, features that are advantageous for low dose or quantitative applications^30–34^. At the same time, these same advances increase reliance on nonlinear processing chains and calibration accuracy, which can introduce systematic errors that are not captured by variance-only metrics. Moreover, while advanced iterative reconstruction techniques improve image quality, they may adversely affect quantitative accuracy^35^. In this context, a fast bias estimator complements CRLB-based design by explicitly quantifying systematic shifts in clinically relevant outputs (e.g., iodine density or virtual monoenergetic HU). This enables acquisition choices that better reflect modern task demands and provide optimal inputs for high-resolution spectral imaging.

## 5. Conclusion

In this work, we present an efficient, practical tool for estimating spectral CT bias. Spectral bias and noise estimator methods are essential for designing accurate quantitative CT systems. As contrast biomarkers become increasingly integrated into clinical and diagnostic workflow, pursuit of low bias acquisitions becomes paramount for spectral CT instrumentation. This estimator provides a computationally efficient complement to existing noise-based optimization methods.

## Data Availability

All data produced in the present study are available upon reasonable request to the authors.

## Acknowledgements

This work was supported in part through grants from the National Institutes of Health (R01EB030494) and Philips Healthcare.

